# Working in a care home during the COVID-19 pandemic: How has the pandemic changed working practices?

**DOI:** 10.1101/2021.06.10.21258611

**Authors:** Clarissa Giebel, Kerry Hanna, Jacqueline Cannon, Justine Shenton, Stephen Mason, Hilary Tetlow, Paul Marlow, Manoj Rajagopal, Mark Gabbay

## Abstract

The aim of this research was to explore the impact of COVID-19 on the working practices of care home staff, caring for people living with dementia. Remote qualitative, semi-structured interviews were conducted with care home staff caring for people living with dementia (PLWD) in the UK. Participants were recruited to the larger programme of research via convenience sampling. Interviews were conducted via telephone or online platforms. This research employed inductive thematic analysis. Sixteen care home staff were included in this study. Three overarching themes were developed from the analysis that conveyed changes to the everyday working practices of the care home workforce and the impact such changes posed to staff wellbeing: (1) Practical implications of working in a care home during the COVID-19 pandemic; (2); Staff values and changes to the staff roles (3): Impact to the care home staff and concerns for the care sector. The COVID-19 pandemic has significantly disrupted the daily working practices of care home staff, with staff forced to adopt additional roles on top of increased workloads to compensate for the loss of external agencies and support. Support and guidance must be offered urgently to inform care home staff on how to best adapt to their new working practices, ensuring that they are adequately trained.

## Introduction

During the pandemic, care homes have experienced significant impacts including visiting restrictions, resident illnesses and consequently, permanent home closures (Martin & Mercer 2020; O’Dowd 2020). By May 2020, care home residents accounted for 54% of all COVID-19 related deaths (Office for National Statistics. 2020a), with dementia and Alzheimer’s disease reportedly the most common pre-existing condition among deaths involving COVID-19 in care homes (Office for National Statistics. 2020b).

Care homes in the UK consist of residential and/or nursing care, with a multi-professional workforce, collectively responsible for the daily health care and personal care of the residents (Perry et al. 2003). Ambiguity around staff roles and responsibilities have been found to exist in the care sector, namely between nurses and care assistants (Perry et al. 2003). The adverse impacts of COVID-19 upon care homes are considerable (White et al. 2021), therefore further challenges and alterations to staff roles are likely to have occurred but are, at present, unreported. There is emerging evidence about the impact of the pandemic on vulnerable groups including those living with dementia, and their unpaid carers (Giebel et al. 2020a; Giebel et al. 2020b), however much less into the impact on care home staffs’ working practices or the impact of the pandemic on the staff, despite heavy media coverage (Cousins, de Vries & Dening 2021). Care home staff are at a greater risk of contracting COVID-19 (Ladhani et al. 2020), particularly if staff are working across multiple care homes (Rios et al. 2020).

Therefore, the aim of this study was to explore the impact of COVID-19 on the working practices of care homes from the perspectives of the care home workforce. Understanding the impact of the pandemic on working practices in institutional long-term care settings could help inform services of areas of change and development required to better support care home staff working through the pandemic, and sustain care homes in the long-term, as there is no guarantee that vaccines and increased testing roll-out will lead to pre-pandemic working practices and visiting rights. This is particularly relevant for lower- and middle-income countries that are struggling to access sufficient vaccine procurement, where the impact of COVID-19 will persist longer.

## Methods

### Participants and recruitment

This study is part of a larger programme of research exploring the impact of the COVID-19 pandemic on care home staff, and family carers with a relative residing in a care home (Ref: 7626).This study describes the responses from care home staff only, in relation to their working practices before and since the pandemic. Care home staff, aged ≥18, who worked in a care home or worked solely with care homes as part of their clinical roles were eligible to take part.

Third sector organisations, many of which have existing links with care home organisations, advertised the study for recruitment. Furthermore, an existing network of dementia and ageing and social media were used to advertise the study. Organisations were contacted and the study information shared with them, whilst information about the study was further posted on social media. Interested participants could contact the principal investigator via email to take part. Ethical approval was obtained through the University of Liverpool ethics committee. An approved participant information sheet was emailed to the participants, and re-read to the participants, prior to taking consent. Verbal informed consent was taken before the interview commenced, which was also audio-recorded, as per the approved ethical protocol.

### Data and data collection

Data were collected between October and November 2020, with different regional restrictions being in place at first, followed by the second England-wide lockdown from the 1^st^ of November until early December. There was no clear guidance in place for care homes during this time.

Semi-structured interviews were conducted, with participants choosing their preferred form of communication (phone or skype/zoom). Interviews were audio-recorded, with verbal consent obtained and recorded at the beginning of each interview. Interviews lasted between 16 and 41 minutes, averaging 24(+/-7) minutes.

A topic guide was developed with clinicians and current and former carers. Participants were initially asked background questions including age, gender, ethnicity, years of education, length of working in the care home sector, job role, and care home size. Staff were asked to discuss their regular working day before and since the pandemic, the viral testing and COVID-19 safety measures employed in their care home, visitations and communications between family members and residents, and the impact of the restrictions on the staff and residents.

### Data analysis

Interviews were transcribed and anonymised prior to coding by research team members (XXX) and one assistant psychologist, who are all experienced in qualitative analysis. The research team consisted of researchers, clinicians, third sector care providers, and included public involvement from former unpaid carers. Each transcript was double-coded blindly by two researchers, and final themes discussed with carers to ensure mutual agreement. Data were analysed using inductive thematic analysis(Braun & Clarke 2006).

### Public involvement

One current and two former unpaid carers were involved in all aspects of the study, including study document design, contribution to group discussion, and interpretation and dissemination of findings. Public involvement fees were paid according to NIHR INVOLVE (2005) guidelines.

## Results

One member of staff from 16 care homes were interviewed. Table 1 summarises their demographic characteristics. In summary, the majority were female (n=13), White British (n=13) and with a mean age of 41.8 years (±16.6). The mean years of working in a care home was 9.3 (±10.6), with the most common job roles being care assistant and manager (n=4 respectively).

**Table 1.**
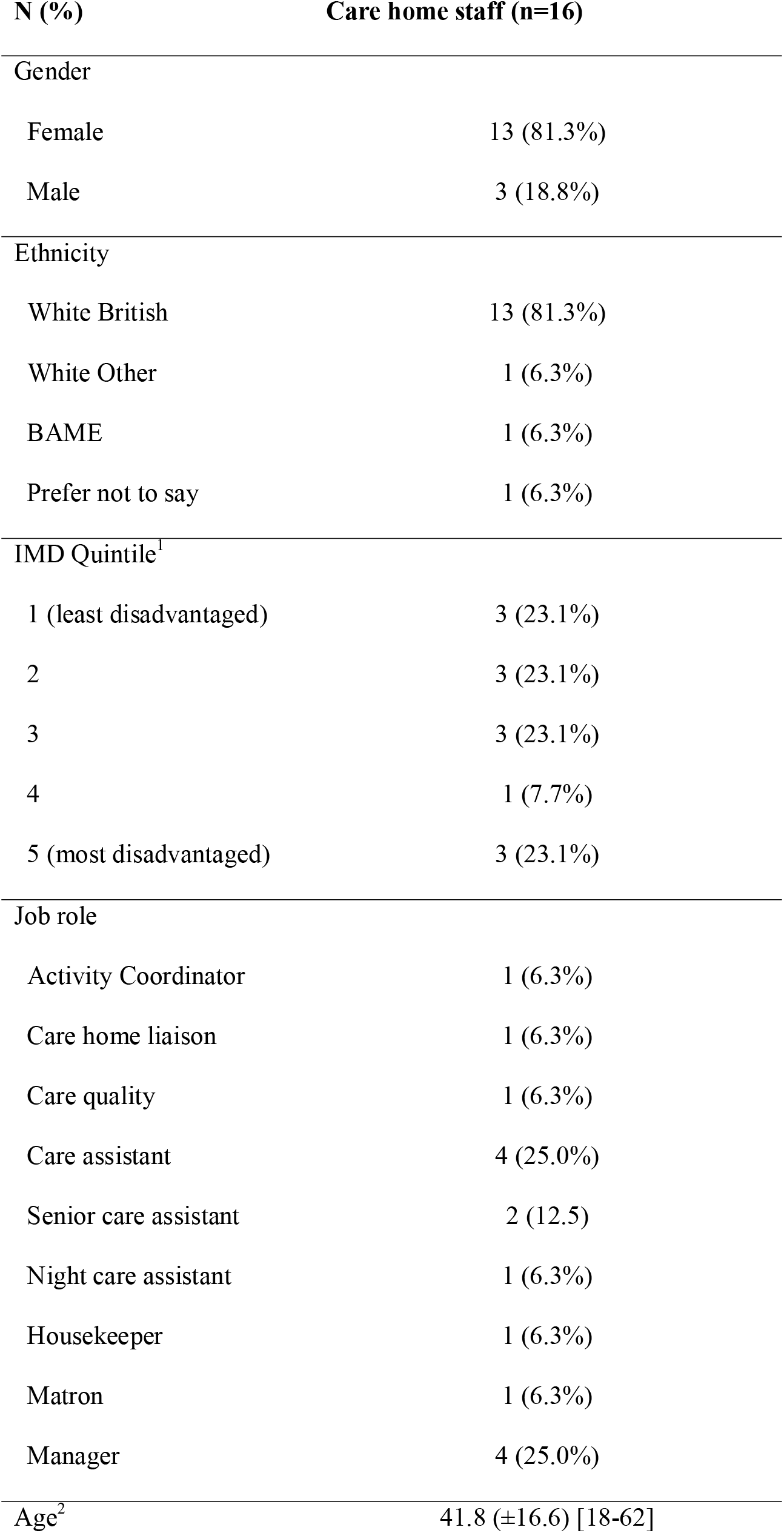
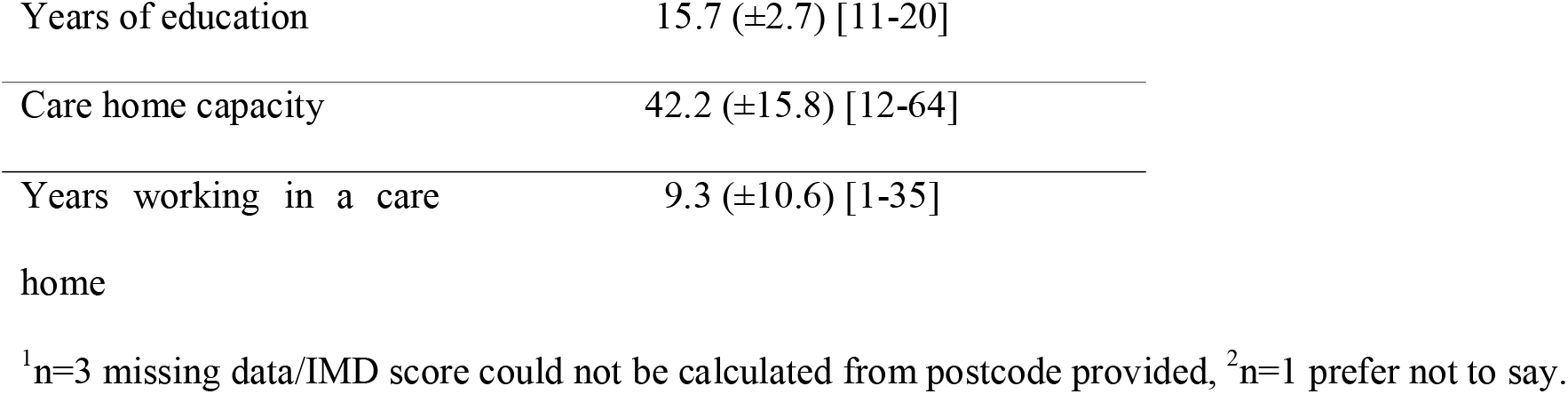
Demographic characteristics of care home staff.

Three central themes with multiple subthemes are described (Table 2): (1) Practical implications of working in a care home during the COVID-19 pandemic; (2); Staff values and changes to their roles (3): Impact to care home staff and concerns for the care sector. Despite recruiting only care home staff working in homes that care for PLWD, participants spoke of their roles in a general sense, and dementia did not present as a significant factor in their narratives around working during the pandemic.

**Table 2.** Coding tree of themes and subthemes identified using thematic analysis.

| Theme | Subtheme |
| --- | --- |
| **1. Practical implications of working in a care home during the COVID-19 pandemic** | Variable and unequal responses to new guidance and restrictions |
|  | Feeling unsupported in complying with new measures |
| **2. Staff values and changes to their roles** | Adopting the role of external agencies |
|  | Adopting a greater emotional/familial role |
| **3. Impact to care home staff and concerns for the care sector** | Increased workload, stress and burnout |
|  | Changes to perception of job |
|  | Concerns over the future of the care sector |

### Theme 1: Practical implications of working in a care home during the COVID-19 pandemic

#### Variable and unequal responses to new guidance and restrictions

Care homes were primarily affected by new public health pandemic protection measures, prompting visiting restrictions and a requirement to comply with new infection control measures. Participants described the initial weeks of lockdown with consensus; no visitors under any circumstances whilst the care homes adapted to the new safety and hygiene guidance.

> *it [the care home] was on immediate lockdown so entertainers, hairdressers, chiropodists, relatives etc. we all of a sudden then had to lockdown our offices so that teams weren’t coming…it’s been quite a lonely, a lonely place the last sort of seven months* **ID41, male care quality manager**

In the weeks that followed, visiting could recommence provided safety measures were put in place. However, staff argued that little central/governmental support was offered to guide them in making the appropriate decisions and adaptations, thus inconsistent visiting abilities were described, suggesting underlying inequalities nationwide.

> *we were allowed to have indoor visits once a week… and then just recently those indoor visits can be extended so they can be in for longer and they don’t have to be physically distancing as long as they’ve got full PPE on, they can have a visitor up to 4 hours …its only once a week* **ID06, female care home manager**

Reasons for this variation often centred on factors that pre-dated the pandemic, outside of the care homes’ control, such as: the location of the care home and the local public health restrictions, the design/setting of the care home in order to accommodate adapted visits (e.g. window visits were possible if residents resided on the ground floor), and sufficient staffing levels to support the new adapted visits. Even where staffing levels were adequate, participants argued that they were not always appropriately trained in supporting the new forms of visitation, such as using new digital platforms, and so these visits were limited.

> *we’ve got this new, it’s a huge iPad on wheels…one resident had a zoom call, I think he enjoyed it…I think we might be encouraged to do it with the residents but I don’t really know… how to set zoom on there so I’d have to be shown how to do that* **ID13, female care assistant**

Staff had to comply with additional infection control measures within their day-to-day roles, including the use of personal protective equipment (PPE) and increased cleaning procedures. However, these measures often caused barriers to their working roles and subsequently added to staff workload.

> *We increased our cleaning procedures… we increased the frequency of cleaning. Day and night … you’re trying to compartmentalise and keep staff separate, we had to take communal areas out because that was a high-risk zone*. **ID18**, male **care home manager**

Variation in the use of PPE was noted, with some homes implementing more stringent protocols than others. A factor in this was the shortage of PPE, especially in the early days of lockdown, and the high cost attached to acquiring available PPE rendering it difficult to purchase in sufficient quantities. Even when participants reported having good supplies of PPE, this was often due to independently proactive managers, donations by local organisations, or at a very high cost. Thus, the participants highlighted a lack of governmental support in obtaining PPE during the pandemic.

> *“we didn’t have enough [PPE], we didn’t have any masks, we had enough aprons and we had enough cloves initially. Then we couldn’t get gloves and we couldn’t get aprons”* **ID04, female housekeeper**

A further area in which staff noted lack of support and variation, was in the testing of staff and residents for the COVID-19 virus. Although testing generally improved over time, protocols for testing varied between homes, and further issues were raised over the reliability of the test results.

> *“we get a test every week now, we didn’t at the start [of the pandemic]but as I say the last, probably about the last two months, we’ve been getting tested once a week”* **ID40, female night care assistant**

#### Feeling unsupported in complying with new measures

Despite instruction, it was not always possible to comply with the new guidance, such as keeping residents socially distanced. In these instances, it was felt that the guidance did not support the needs of the residents, and the staff were not confident in making the appropriate changes. Therefore, adapted visits were often the result of staff innovation, at times even going against government guidelines.

> *“the guidelines changed so much to the point that it was like everyday new guidelines, sometimes twice a day it would change”* **ID38, female care assistant**

At the centre of discussion around support with the new health guidance, staff referred to good and bad management, and how this influenced their compliance. Despite the hardships of working under the strained conditions of the pandemic, staff appreciated the example set by care home managers who supported frontline care during the pandemic.

> *I did think about working from home for some time but then I thought about the team I support… I saw that the staff were quite appreciative of the fact that, as a manager, if I can come in then they can come in as well* **ID37, female care and compliance manager**

In contrast, poor management was described where staff felt the job role had changed significantly, and they were now being asked to do more than was feasible. In some cases, staff reported leaving their job role altogether due to this lack of support.

> *those care homes that had coronavirus at the beginning were not supported. It was an oversight from*
>
> *Governments, there should have been a - pronged approach…care homes have vulnerable people in*
>
> *them… those care homes didn’t have PPE, they weren’t able to get PPE and yet everything was*
>
> *focussed on the NHS…* **ID06, female care home manager**

### Theme 2: Staff values and changes to their roles

#### Adopting the role of external agencies

Following the loss of external agencies and family carers, staff were left to fill the roles that added to the residents’ holistic care. Staff described innovative person-centred approaches that focussed on protecting staff and residents, and often required good relationships with family carers for support.

> *Our staff actually are doing more and more activities on a one to one with people so they’re getting more time spent with them so really, they’re the surprise in all of this, because they have adapted really well* **ID16 care home manager**

However, the loss of entertainers and family visits that punctuated significant portions of the residents’ days, designed to engage them in social interaction and activities, meant that staff where now responsible in suggesting and executing innovative ways to keep residents entertained and morale raised.

> *it was hard as well trying to find activities to do every single day in the afternoon where they’d normally have the family there…people don’t want to do the same thing so that made it difficult* **ID09 female care assistant**

These approaches required adaptations that questioned the suitability of the government’s guidelines within care homes. Staff noted a difference between patient care and patient safety when care conflicted with the recommended guidance, in instances where this was available, leaving them to make difficult management choices.

> *Visitors weren’t allowed to come… [but] we had a room…at the very end of the building, you could access it from outside so that [visitors] didn’t have to walk through the home…I got a call from the Council saying this is contrary to what Public Health England are saying, you must stop it, and they were reporting me to Public Health England* **ID25 female nurse matron**

#### Adopting a greater emotional/familial role

With no visitors, staff found themselves adopting familial roles to support the residents’ emotional wellbeing. However, with residents frequently concerned that their family had stopped visiting, comforting them became challenging. Furthermore, when family telephoned enquiring about their relative, care home staff felt ill prepared for dealing with these deteriorations; unsure whether or not it was ethical to withhold distressing information from the family.

> *We can give hugs and touches but we’re not their family as much as we try to be we can’t replace their family*. **ID32, female activity coordinator**

### Theme 3: Impact to care home staff and concerns for the care sector

#### Increased workload, stress and burnout

Following the aforementioned changes to staff’s job roles and a reported lack of support in following new regulations, the care home staff described higher levels of work-related stress and burnout, exacerbated by staff shortages where staff had to isolate due to the virus.

> *it was like being lost at sea, actually that’s how it felt …my working week went from, well its normally about 50 hours to probably 70… so just generally the day became a lot harder and the anxiety and stress became a lot harder*. **ID06, female home manager**

Furthermore, personal issues due to the strain of public health measures affecting life outside of work was an occurrence for many staff due to the pandemic, including isolation in lockdown, family illness and the loss of normal stress relieving activities, which intensified work-related issues.

> *Personal stress is an issue…*.*my management of my stress was going to the gym and just winding down, but of course that can’t happen …I’ve talked to managers and deputy managers from other homes and it’s pretty similar, the stress doesn’t stop* **ID18, male care home manager**

The underlying anxiety of contracting and/or transmitting the COVID-19 virus, and the effect this would have on oneself and those around them, was a prevailing point of concern for many staff.

> *…I contracted COVID myself and then I was off for couple of months…I was in tears, I had loads of negative thoughts…when I was going back to work I was really feeling nervous and lack of confidence and thinking oh my God what if something again happens to me and how will I survive and what will I do and how will I provide the care* **ID36, female senior care assistant**

#### Changes to perception of job

As the pandemic created unexpected changes to the care home staff roles, this led them to consider how they perceived their jobs, both positively and negatively, in comparison to the time before COVID-19. Staff expressed a loss of enjoyment in their work due to the newfound stress they found themselves working under, namely an increased workload, the demand on infection prevention superseding usual patient care, and the observed impact on the residents’ wellbeing.

> *I dislike my job intensely…I wanted to walk away from my job in June. I was incredibly stressed and felt anxious continually. My GP actually was phoning me once a week… I personally was at the point where I was like I don’t want to do this job anymore because it’s just not a job that’s doable* **ID06, female care home manager**

Others reported a positive perception of their job role from working during the pandemic. These staff conveyed a sense of pride, stemming from their role of caring for some of the most COVID-vulnerable in society. Nevertheless, it was still clear that care home staff were viewing their roles in a different light, due to the extreme pressure and that they had to work through.

*I do love my job, I suppose it’s made me see it different, I think it’s been hard, it’s been harder…my sister and my daughter are constantly like [we are] really worried about you* **ID40, female night care assistant**

#### Concerns over the future of the care sector

Following negative media reports of high numbers of COVID-related deaths in care homes and restrictions on visits, staff raised concerns over the growing stigma of care homes, and the impact this will have on attracting future residents.

> *the problem is that care homes got such bad press at the beginning of COVID that I think a lot of care homes are frightened to let relatives in, in case COVID comes in and then they get the blame*… **ID06, female care home manager**

Furthermore, due to significant losses in care home beds due to COVID-19 related deaths, homes now faced financial repercussions and closures. In addition, the lower number of residents resulted in higher levels of overstaffing. Staff reported fears that some will lose jobs or be relocated to other care homes as a result of this, adding to their previous feelings of work-related stress.

> *…at the worst we had ten empty beds which probably [in a] 62 bed home doesn’t sound much but*
>
> *you’re talking about a lot of money every month and if we were private, we might have decided to cut*
>
> *our losses and close or sell up as a number of homes around here have* **ID18, male care home manager**

In addition, staff reported fears for the future of the care sector in general, postulating that fewer people will seek to work in this field due to perceptions of lack of support and high levels of work pressure.

> *I don’t feel like people will want to work in care after this, I think it’s frightened a lot of people off* **ID09, female care assistant**

Discrepancies between the government’s responses to support the NHS versus the care sector were frequently highlighted, concluding that the care sector was comparatively neglected. Overall, the care home staff expressed a desire for their roles, and the care sector in general, to be recognised for the work they have, and are continuing to, contribute to the fight against COVID-19.

## Discussion

This study is one of the first to report the impact that the COVID-19 pandemic is having on the care home workforce. Impacts include the high risk of staff catching and transmitting the coronavirus, changes to staff job roles, and subsequent stress placed upon staff in response to the pandemic’s public health measures.

Care home staff reported feeling unsafe and unprotected from the virus whilst working during this time, originating from initial PPE shortages. The lack of PPE for care home workers has been reported in the literature and media (Gordon et al. 2020), however, the current study has further explored the emotional and physical impact that this shortage caused to staff. Our findings suggest that in the short term, staff were forced to adapt to the circumstances, accepting donations or reusing PPE, whilst spending large amounts of money to procure supplies. However, in order to address PPE shortages longer-term, or in the event of further pandemics, better integration of procurement and supply is required (Gibson & Greene 2020; Gordon et al. 2020).

Furthermore, regular COVID-19 testing of both the staff and residents varied, supporting earlier research findings where staff shared concerns that the results of residents’ tests were unreliable (Nyashanu, Pfende & Ekpenyong 2020), especially in instances where the resident was difficult to swab due to cognitive issues, namely PLWD. Recent evidence suggests regular testing that produces rapid and reliable results is highly important in this high-risk population of care home residents, as infections spread quickly, and presentations are often atypical (Gordon et al. 2020). However, the current study provides additional evidence highlighting the safety issues that staff are consequently facing at work when testing is inconsistent and/or unreliable. With recent evidence identifying care home staff at an increased risk of contracting COVID-19 (Ladhani et al. 2020), it is imperative that issues around staff and resident testing are addressed.

This research identified that the loss of external agencies and family carers, through visiting restrictions, impacted both staff and residents. Before the pandemic, family members remained active carers, evidently supporting care home staff in delivering levels of personal care, emotional support and providing residents with a link to the outside world (Davies & Nolan 2006). Without the input of family carers, amongst other external roles, staff reported being overworked and unsupported. In Italy, healthcare staff treating COVID-19 patients further conveyed similar emotions of stress and burnout (Nyashanu, Pfende & Ekpenyong 2020; Trumello et al. 2020), however, the current study adds to this evidence base, exemplifying the similar impact that care home staff experience when caring for vulnerable residents, and observing the negative effects of the virus first hand.

The findings reveal a stigma of care homes with high infection rates and a burnt-out staff force that has emerged through the time of COVID-19. The study identified a reduced enjoyment of job roles amongst participants, due to the stressful conditions under which they have been working, supporting earlier reports that care home staff have felt devalued and unsupported when working in the time of COVID-19 (McGilton et al. 2020). For families to feel confident in care homes, and staff to feel valued in their roles, the public image of care homes must be altered. However, this can only be done by supporting the care homes during the pandemic at government level, through providing them with sufficient resources and funding (McGilton et al. 2020).

Overall, it is apparent from this research that staff roles have changed significantly and unexpectedly during the pandemic with little or no support offered to guide staff role transition. Despite the demands of the job, care assistants in the UK require only entry level qualifications (National Careers Service. 2020), the lowest level of national qualification requirement. Participants acknowledged that their skillset must now change to accommodate the role alterations arising from the pandemic, which concurs with earlier reports that care staff require additional support to undertake their roles safely since the time of COVID-19 (Fallon et al. 2020). Both existing and new staff require appropriate training on infection control and adapting to new roles and responsibilities. With previous issues noted around care home staff retention and turnover rates generally in the UK (Costello et al. 2019; Donoghue 2009), ensuring that standardised training is undertaken may be challenging and must be considered in future training and service planning.

There are some limitations to note. Fewer participants from BAME backgrounds undertook this research due to the convenience sampling method, and future research strategies should consider an alternative method in order to capture the views of a broader population. The authors acknowledge that interview quality may be limited if not conducted face-to-face, however the research did not observe any issues during the interview process.

## Conclusion and implications

This research has identified drastic changes to care home staff roles during the pandemic. However, staff concerns expand beyond individual impacts of the pandemic, and consider the wider impact on the future of care homes. The findings from this study highlight a gap in support and guidance that must be addressed urgently to inform care home staff on how to best adapt to their new working practices, ensuring that they are adequately trained. Such measures will lend to protecting staffs’ health and wellbeing, and ensuring care homes sustainably recover from the pandemic long-tem.

## Data Availability

The data that support the findings of this study are available on request from the author [CG]. The data are not publicly available due to ethical restrictions.

## Declaration

The authors have no conflicts of interest to declare.

## Acknowledgements

We wish to thank all participants in taking part in this study. We also wish to thank Thomas Faulkner for helping with the analysis of some transcripts, the organisations who have helped with recruiting and/or analysing data, and Maxine Martin and Lynn McClymont for transcribing the audio files swiftly.

